# Recombinant zoster vaccination in patients with dementia is associated with improved survival and better cognitive preservation

**DOI:** 10.64898/2026.04.09.26350509

**Authors:** Katarina Soltys, Roni Sara-Buchbut, Noga Ish Shalom, Joshua Stokar, Benjamin Y Klein, Ronit Calderon-Margalit, Charles L Greenblatt, Moshe Shay Ben-Haim

## Abstract

Dementia affects tens of millions of people worldwide, yet disease-modifying treatments remain strikingly limited. Although the recombinant zoster vaccine Shingrix has been associated with reduced dementia incidence, its potential influence on individuals already living with dementia is unknown. Here, we followed a propensity-score matched cohort of 68,960 US dementia patients using a nationwide electronic health record network, comparing Shingrix recipients within two years of diagnosis to recipients of any other vaccine. Shingrix was associated with substantially reduced all-cause mortality across the first three years of follow-up (hazard ratios 0.74, 0.88, and 0.89; P≤0.006), robust across multiple sensitivity analyses. Furthermore, within-individual subgroup analyses of repeated Mini-Mental State Examinations conducted 3–6 years apart revealed significantly divergent cognitive decline rates across groups (time-by-group interaction P=0.002). Interval vaccination was associated with more stable cognition, contrasting with steeper declines in unvaccinated individuals. These findings support prospective evaluation of recombinant zoster vaccination as a potential strategy to improve outcomes in patients with established dementia.

## Introduction

Dementia is among the most consequential disorders of later life, imposing a steadily growing burden of disability, dependency and death worldwide. In 2021, an estimated 57 million people were living with dementia, leading the World Health Organization to recognize dementia as a major cause of disability among older adults and the seventh leading cause of death globally^1,2^. Despite this clinical and societal burden, the therapeutic options for people already living with dementia remain strikingly limited. For example, anti-amyloid therapies in Alzheimer’s disease have shown modest, if any, effect in modification of the course of disease, and their high cost further limits their use^3,4^. There is therefore an urgent need to identify interventions that can attenuate or slow down disease progression.

One increasingly important line of inquiry concerns the interface between the immune system and neurodegeneration in old age. A large body of research suggests that chronic inflammation, dysregulated innate immunity, microglial activation and neuro-immune signaling may contribute to the development and progression of dementia^5–8^. In parallel, neurotropic pathogens, including members of the herpesvirus family have been proposed as potential contributors to neurodegeneration over time^9–12^. Varicella zoster virus (VZV) is particularly relevant in this context. Following primary infection, it can remain latent for decades, and its reactivation, which becomes more common in aging, has been linked to vasculopathy^13^, stroke^14^ and dementia^15^.

More recently, observational and quasi experimental work has suggested that VZV vaccination is associated with a lower risk of subsequent dementia^16,17^, an association that was also observed with other vaccines^18–21^. Natural experiments that exploited policy-based eligibility thresholds helped address healthy-vaccine biases, and suggested that zoster vaccination might reduce dementia related outcomes later in life ^22, 23^. For example, a recent study harnessed a rapid shift in vaccine type uptake after the introduction of the recombinant zoster vaccine (Shingrix) in the United States and reported that individuals receiving the new recombinant vaccine (91.3% efficacy in herpes zoster prevention in > 70 year-olds^24^), had a lower risk of incident dementia compared with individuals receiving the older live-attenuated vaccine Zostavax^22^ (that has a lower efficacy of 37.6% in herpes zoster prevention in >70 year-olds^25^). A more recent study has investigated individuals who were already diagnosed with dementia and found that the Zostavax vaccine was associated with reduced mortality compared to those who were not eligible, based on birth date eligibility thresholds^23^. This latter study raises a provocative possibility that the clinical course of dementia may remain modifiable even after diagnosis, and that a widely used adult vaccine could influence outcomes far beyond its original infectious disease indication. Nevertheless, this study was based on the discontinued Zostavax vaccine and from a relatively small cohort. Current practice has shifted to Shingrix, a recombinant adjuvanted vaccine containing herpes zoster glycoprotein E and the AS01B adjuvant system^26^, and Zostavax is no longer available in the United States^27^. This is critical, since the recombinant and live attenuated vaccines differ fundamentally in composition, efficacy, immunogenicity and the ways they engage with immune programs. It therefore cannot be assumed that responses observed with older vaccines are unconditionally generalizable to the recombinant vaccine that clinicians currently use.

Here, we tested whether the new recombinant Shingrix vaccine administered to individuals already living with dementia is associated with improved survival in a substantially large real-world cohort derived from a US-based network of electronic health records. Notably, to account for healthy vaccine bias that is often associated with observational studies, we compared Shingrix vaccinated with a group of dementia patients receiving any vaccine other than Shingrix and also applied tight propensity score matching to control for numerous potential socio-demographic, socio-economic confounders and prior comorbidities. Furthermore, to test whether vaccination is associated with better cognitive preservation over time, we examined a subset of patients with available repeated Mini-Mental State Examination (MMSE) cognition scores for their individual rate of cognitive decline across a 3-6 year period.

## Results

### Shingrix vaccination after dementia diagnosis is associated with lower all-cause mortality

We assembled a retrospective cohort of patients with a recorded diagnosis of dementia and identified those who received Shingrix within two years after diagnosis. Within this population, 34,543 patients met exposure criteria for Shingrix and 200,176 met comparator criteria as recipients of any other vaccine except Shingrix. After 1:1 propensity score matching, our study population included 68,960 individuals (34,480 individuals in the Shingrix group and 34,480 patients in the comparator group, with good balance and standardized mean difference ≤0.041; Supplementary Table 1).

To avoid immortal time bias, survival follow-up began only after completion of the 2-year post-diagnosis vaccination window and continued for up to 5 years thereafter. Kaplan-Meier curves separated early and remained apart over time, consistent with a survival advantage among Shingrix recipients (Fig.1a). While Shingrix vaccination was directionally associated with a reduced risk of all-cause mortality (HR=0.86,95% CI:0.83-0.89; P<0.0001), the proportionality assumption did not hold (P<0.0001). We therefore stratified the analysis in year-long follow-up windows in which the proportionality assumption was not violated (P≥0.23). The association was the strongest early at the 1^st^ year with a hazard ratio of 0.74 (95% CI:0.70-0.79; P<0.0001). It remained significant at year 2, with a hazard ratio of 0.88 (95% CI:0.82-0.95; P=0.0005), and at year 3, with a hazard ratio of 0.89 (95% CI:0.82-0.97; P=0.006). By years 4 and 5, the point estimates had attenuated toward the null, with hazard ratios of 0.98 (95% CI:0.89-1.07; P=0.61) and 0.995 (95% CI:0.90-1.11; P=0.92), respectively.

**Figure 1.**
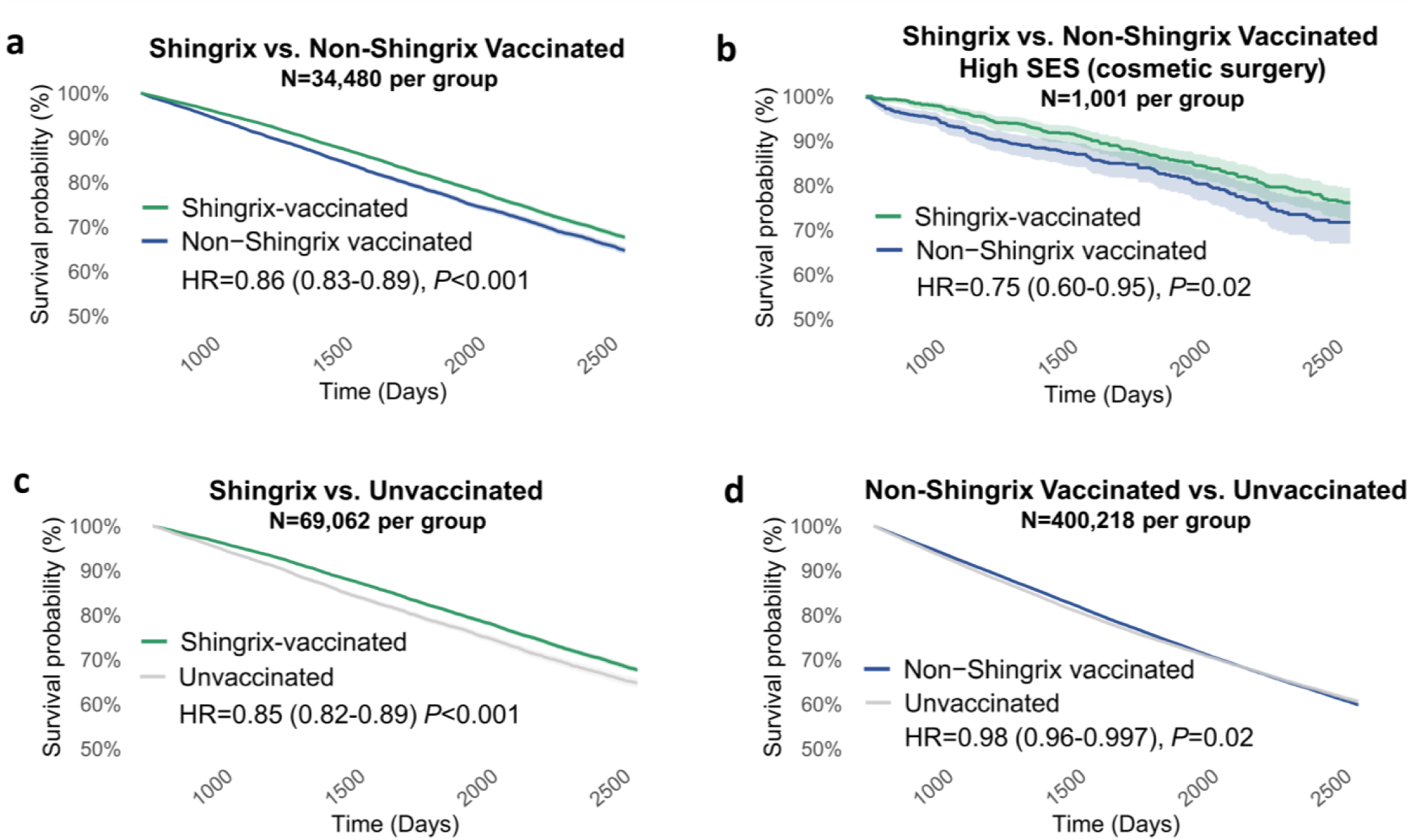
Survival according to vaccination status 2 years after dementia diagnosis. Kaplan-Meier curves show post-diagnosis survival probabilities in patients with dementia across vaccination groups. **a.** Shingrix-vaccinated patients compared with non-Shingrix-vaccinated patients in the primary matched analysis. **b.** Shingrix-vaccinated patients compared with matched non-Shingrix- vaccinated patients in a subgroup with high socioeconomic status, proxied by a history of cosmetic surgery. **c.** Shingrix-vaccinated patients compared with matched unvaccinated patients. **d.** Non-Shingrix-vaccinated patients compared with matched unvaccinated patients. Hazard ratios (95% confidence intervals), and log-rank P values are shown in each panel. Shaded areas around the lines indicate 95% confidence intervals. HRs below 1 indicate lower mortality in the first-named group.

To gain insight on the magnitude of the association relative to unvaccinated individuals, we compared Shingrix vaccinated with unvaccinated matched dementia patients (N=69,062 after matching). The HR was 0.85 (95% CI:0.82-0.89; P<0.0001), see Fig.1c. In contrast, the group receiving any vaccine other than Shingrix showed a much smaller benefit over the unvaccinated (N=400,218 after matching, HR=0.98, 95% CI:0.959-0.997; P=0.0234, Fig.1d).

### Sensitivity analyses across different subgroups and by zoster history

We further examined whether the association was modified in a stratification across different relevant subgroups (see Fig. 2). The association was significant in both women and men, across age strata, across vaccination timing, and dosages. Moreover, when restricting the cohorts to specific dementia types, or race categories, we observed similarly significant HRs (except in dementia with Lewy bodies, and in native Hawaiian or other Pacific Islander subgroups that had considerably lower N, Fig.2). Furthermore, because protection against varicella-zoster reactivation might plausibly contribute to improved outcomes, we also examined whether the association depended on herpes zoster infection. Here, the signal remained evident even among individuals who never had a recorded history of shingles or chicken-pox, and was also relatively similar in individuals that did have shingles or chicken pox recorded in EHR (Fig.2).

**Figure 2.**
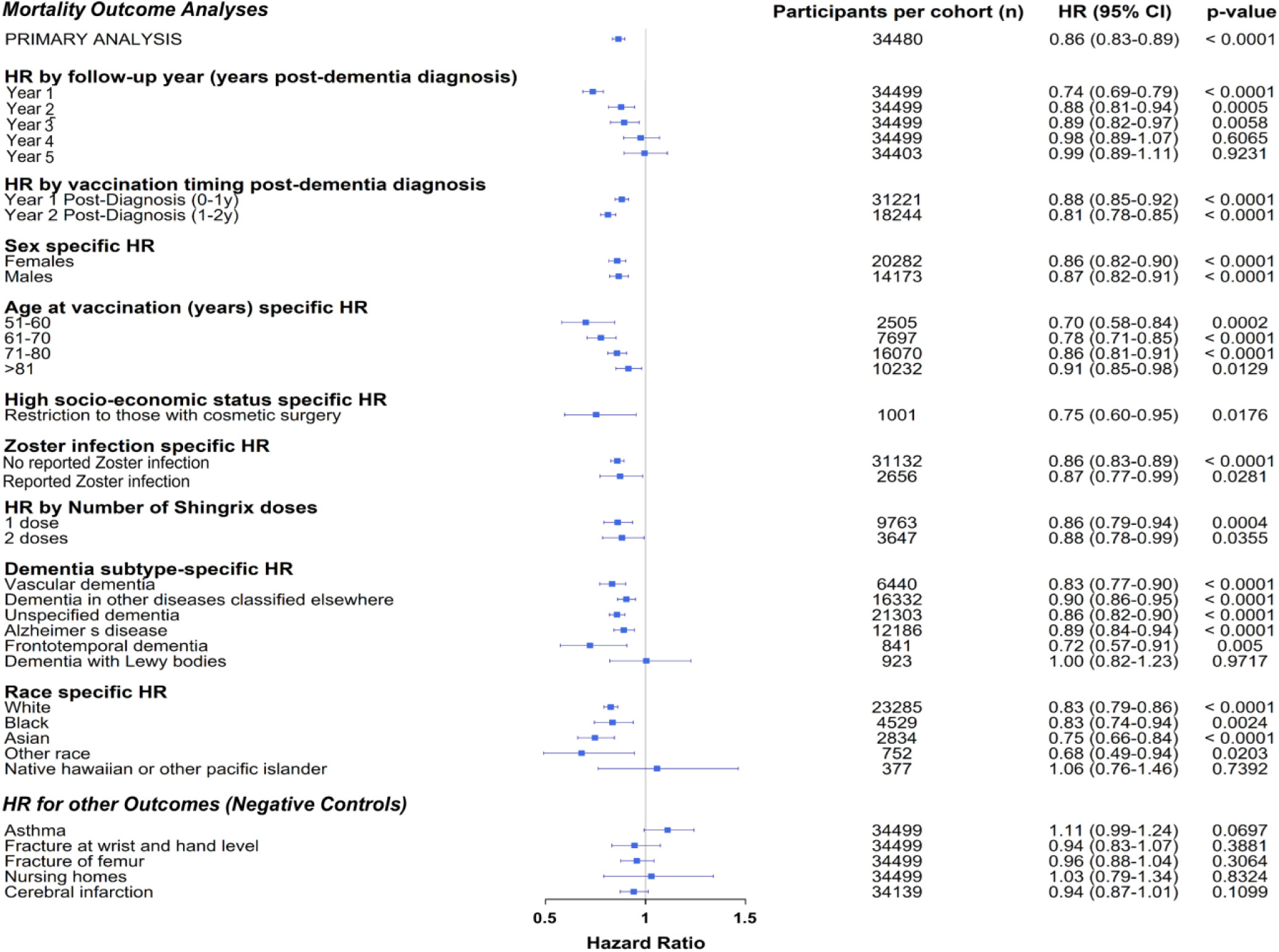
Association of Shingrix vaccination vs. matched non-Shingrix vaccinated across strata. Forest plot showing hazard ratios (HRs), 95% confidence intervals, and the number of matched dementia patients for all-cause mortality associated with Shingrix vaccination compared with matched non-Shingrix-vaccinated patients across prespecified strata. Negative-control outcomes include non-mortality outcomes assumed to be less affected by Shingrix vaccination. HRs below 1 indicate lower mortality in the Shingrix-vaccinated group.

Since residual confounding by care access and socioeconomic structure are among the most plausible threats to interpretation, we performed an additional subgroup analysis of dementia patients with a recorded history of cosmetic surgery as a proxy for higher healthcare access and elective care use. Though the N was much lower (N=1,001 matched patients per cohort), the association remained significant, with an HR of 0.75 (95% CI:0.60-0.95; P=0.018, Fig.1b, Supplementary Table2).

Finally, to test whether Shingrix exposure is not merely reflecting a generalized tendency towards lower coded risk across any outcome, we examined negative-control outcomes in which Shingrix vaccination may have little or no influence. In analyses following asthma as an outcome, or that of fractures of wrist, or femur fractures, or cerebral infarction, or nursing home residence (proxy for institutionalization), the associations were not statistically significant (Fig.2). These near-null findings do not exclude all residual confounding, but they argue against a simplistic explanation in which Shingrix recipients appear systematically protected against any and every outcome.

Furthermore, to ensure that the main results were not driven by matching, we repeated the analysis in the unmatched original cohorts. Across a series of progressively adjusted Cox models, from an unadjusted model to models incorporating sociodemographic, lifestyle, health-seeking, socioeconomic, and clinical factors, the association remained highly significant, with HRs ≤ 0.82 (P<0.0001; Fig. 3; see also Supplementary Table 3 for the full model 6), supporting the robustness of the findings.

**Figure 3.**
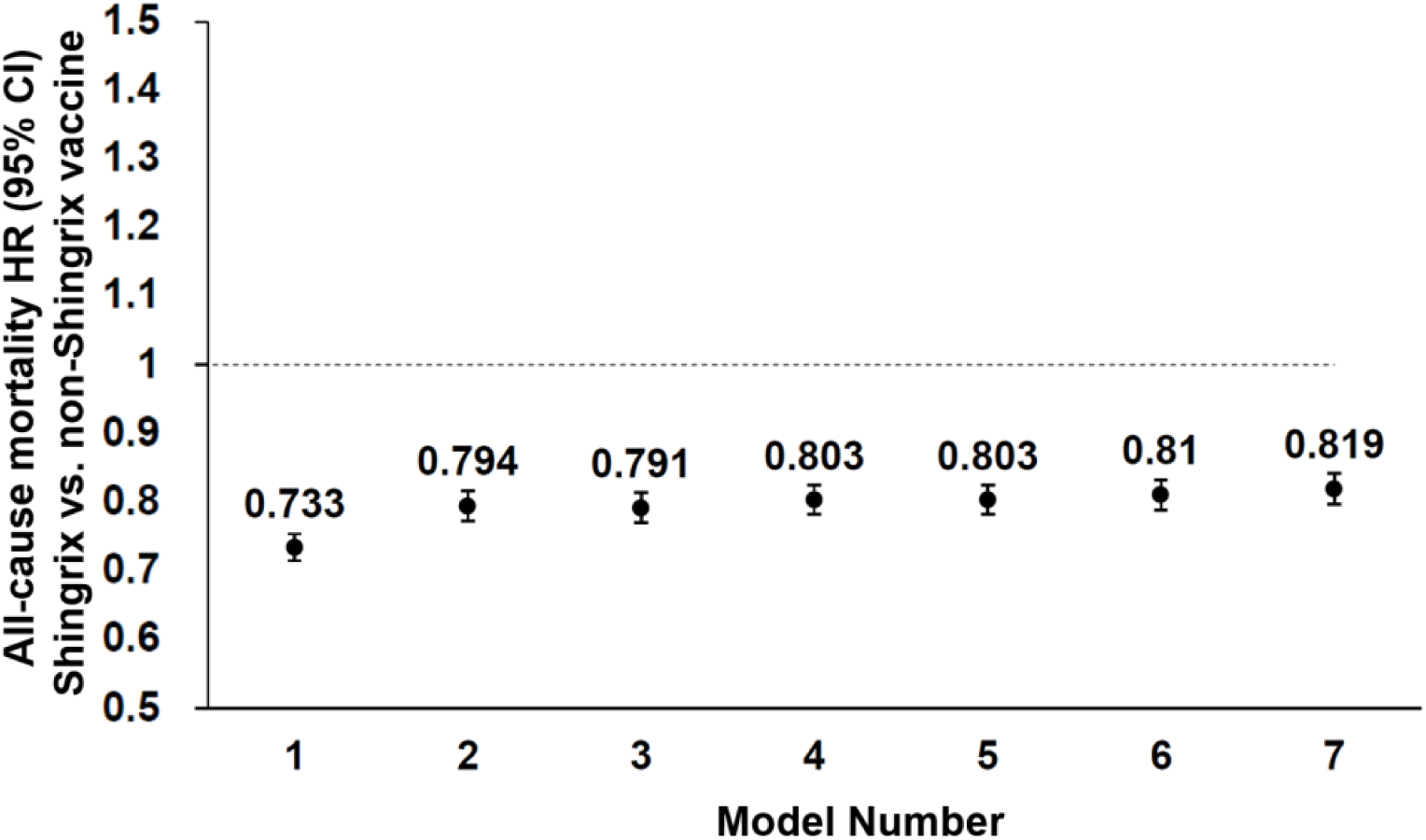
Robustness of the association between Shingrix vaccination and mortality across progressively adjusted Cox models. Forest plot showing hazard ratios, HRs, and 95% confidence intervals for all-cause mortality comparing unmatched Shingrix-vaccinated (N=34,543) with non-Shingrix-vaccinated patients (N=200,176) across a series of Cox proportional hazards models with sequential covariate adjustment. Models were adjusted cumulatively starting from a crude model without covariates (model 1). Model 2 included sociodemographic factors (age at index, sex, race); Model 3 included also lifestyle factors (BMI, smoking, alcohol abuse); Model 4 included also health-seeking behavior (general check-ups, long term medication use, cancer screening). Model 5 included also socioeconomic determinants of health (employment, housing, health insurance, private funding). The full model (model 6) included also selected comorbidities (diabetes, cerebrovascular diseases, cardiovascular diseases, Parkinson’s disease, liver cirrhosis, kidney disease, MMSE, and blood pressure) see also Supplementary Table 3. Model 7 included an expanded set of additional 43 comorbidities based on a recent study^22^, as detailed in Supplementary Table 4.

### Repeated MMSE measurements support better cognitive preservation in vaccinated individuals

To explore whether survival was accompanied by differences in cognitive trajectory, we identified in the network a subgroup of patients who underwent repeated Mini-Mental State Examination (MMSE) cognition scores assessed 3 to 6 years apart, classifying them according to their vaccination status in the interval between the two measurements. This within-group analysis, comparing early and late cognitive performance of the same patients, substantially reduces residual confounding by remaining unmatched factors and group differences. Among unvaccinated individuals (N=214), the mean MMSE score dropped from a mean of 27.17 (SE=0.27) to 25.77 (SE=0.424), P=0.006. By contrast, patients who received the Shingrix vaccine between the two measurements (N=137) did not experience a noticeable decline, with means shifting negligibly from 28.11 (SE=0.21) to 27.92 (SE=0.26), P=0.572. Similarly, patients who received any non-Shingrix vaccine (N=1,228) also showed stable cognitive trajectories, with mean scores moving from 28.36 (SE=0.08) to 28.30 (SE=0.10), P=0.675; Fig. 4a. Notably, the overall rate of cognitive decline differed significantly across the cohorts (Time × Group interaction P=0.002).

**Fig.4.**
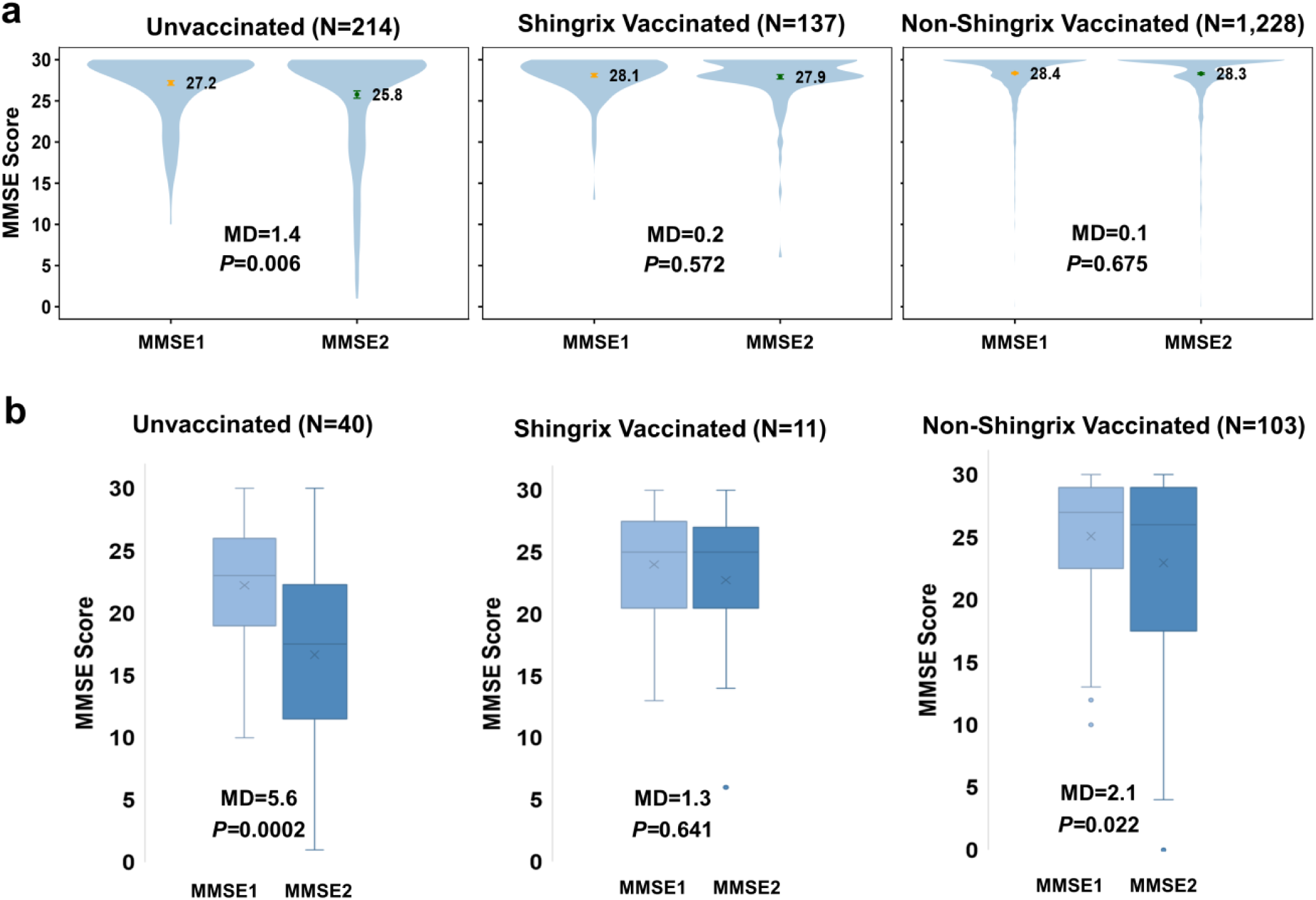
Repeated MMSE measurements support better cognitive preservation in vaccinated individuals. **a.** Violin plots show the distribution of Mini-Mental State Examination (MMSE) cognition scores at the first (MMSE1) and second (MMSE2) measurements, obtained 3 to 6 years apart, in the broader cohort without restriction to recorded dementia. **b.** Boxplots show the corresponding distributions in the subgroup of individuals with recorded dementia. In both panels, patients were grouped according to vaccination status during the interval between the two measurements: unvaccinated, Shingrix-vaccinated, and vaccinated with non-Shingrix vaccines. Numbers above each panel indicate sample size. In panel a, colored markers indicate group means, with error bars representing standard errors. In panel b, boxes represent interquartile ranges, center lines indicate medians, crosses indicate means, and whiskers extend to the most extreme non-outlier values within 1.5 times the interquartile range; points beyond the whiskers are shown as outliers. Mean differences (MD) between MMSE1 and MMSE2 and two-tailed P values are shown in each panel.

Given that this broad group of patients had relatively high starting MMSE scores, we next examined a smaller group of patients that also had a recorded diagnosis of dementia. In this subgroup analysis too, unvaccinated dementia patients (N=40), had a substantially larger decline from 22.23 (SE=0.72) at the first assessment to 16.65 (SE=1.2) at the second assessment (Fig. 4 left panel; P=0.0002). Individuals with dementia who received any vaccine other than Shingrix (N=103) showed a more modest, yet significant, decline from a mean of 25.09 (SE=0.52) to 22.97 (SE=0.76) (P=0.022). Notably, the subset of dementia patients with recorded Shingrix vaccination between the two measurements (N=11) experienced the smallest drop from a mean of 24.00 (SE=1.49) to 22.73 (SE=2.09) (Fig. 4b; P=0.641). Although the formal three-group interaction was likely underpowered due to the small Shingrix sample size (Time × Group interaction P=0.11), pooling the vaccinated cohorts to overcome this limitation revealed that interval vaccination was associated with a significantly attenuated rate of cognitive decline compared to the unvaccinated dementia paitents (Time × Group interaction P=0.037).

These findings, combined with the association observed in the large propensity-matched patients, support the possibility that Shingrix vaccination in dementia patients may be associated not only with lower mortality, but also with better preservation of global cognition. Given the limited disease-modifying options currently available for dementia, together with the availability and established use of Shingrix, these findings justify serious prospective evaluation.

## Discussion

In our large propensity score matched cohort of 68,960 patients, receipt of Shingrix after dementia diagnosis was associated with reduced all-cause mortality compared with receipt of other vaccines. This association persisted across multiple sensitivity analyses, including analyses addressing healthcare access, socioeconomic status, and other sources of bias. Furthermore, the comparison with a broad group vaccinated with any vaccine other than Shingrix argues against the most common threat of healthy-vaccine bias that can exist in many observational studies with only an unvaccinated comparison group.

The longitudinal analyses of repeated cognition scores with the Mini Mental State Examination (MMSE), one of the most widely used tests to screen for cognitive impairment by clinicians^28^, provided supportive evidence that interval vaccination was associated with slower cognitive decline. Although only a subset of patients had two MMSE measurements 3 to 6 years apart, the broader analysis included at least 137 individuals per group and compared patients with their own earlier performance, reducing susceptibility to residual between-group confounding. In that sense, the MMSE results strongly address a central interpretive challenge in observational vaccine research, and provide compelling concordant results to the mortality findings. Importantly, the cognition results are also concordant with recent observations showing trends for improved cognitive preservation in individuals receiving the BCG tuberculosis vaccine^29,30^.

While most prior work focused on delaying dementia onset^16,17,22^, our study examines whether vaccination with the newer Shingrix vaccine may influence outcomes even after dementia has already been manifested. Although the older live attenuated vaccine, Zostavax, has recently shown possible survival benefit in a small cohort of dementia patients^23^, these results could not simply be extended to the newer recombinant adjuvanted vaccine that is currently used in clinical practice, which differs fundamentally from Zostavax in formulation, efficacy, and immunological effects. Patients with established dementia are a particularly vulnerable population, and even modest improvements in survival or cognitive preservation could have substantial clinical and public health implications.

If the effects are truly beneficial, several non-mutually exclusive mechanisms could plausibly explain these associations. One possibility is that prevention of varicella-zoster virus reactivation reduces downstream neurologic, inflammatory, and vascular insults that may accelerate decline in vulnerable elderly patients ^9–12, 15^. Another is that Shingrix exerts effects beyond prevention of clinically apparent shingles. More generally, a growing number of studies on vaccine off-target effects and trained immunity suggest that vaccines may alter inflammatory responses beyond pathogen-specific protection^31–33^. Unlike Zostavax, Shingrix is a recombinant glycoprotein E vaccine formulated with the AS01B adjuvant, designed to induce strong cellular and humoral immune responses in older adults. In our results, restricting the analysis to individuals with documented zoster/chicken pox, or only to those without documentation, yielded similar protective hazard ratios. It is therefore plausible that the association reflects not only pathogen-specific protection but also broader immune effects relevant to dementia progression. Our study, therefore, cannot distinguish between these possibilities, but it strongly suggests that the clinical implications of recombinant zoster vaccination in dementia warrant further investigation.

Notably, the study has some limitations that should be taken into account. Primarily, the mortality associations are still observational and cannot therefore establish clear causality. Propensity score matching did balance the most relevant measured covariates and comorbidities well, but not unmeasured or imperfectly measured factors. Disease severity, and some socioeconomic factors such as education level are incompletely captured in EHR data. Vaccination history and clinical care received outside participating health organizations may also have been incompletely recorded. These considerations should therefore be taken into account when interpreting the association findings. Despite these limitations, it is noteworthy that the associations were robust across all available relevant covariates, and multiple sensitivity analyses designed to address potential threats to validity in a substantially large group of dementia patients with a broad active comparator group of individuals vaccinated with any other vaccine. Furthermore, the convergence of the mortality and MMSE findings is striking. The repeated MMSE analysis is also less vulnerable to residual confounding from between-patient differences. Together, these findings strengthen the possibility that the association reflects more than a general healthy vaccinee effect and raise the possibility that recombinant zoster vaccination could influence clinically meaningful outcomes even after dementia has manifested.

Taken together, our findings suggest that recombinant zoster vaccination may be associated with clinically meaningful benefit even after dementia has already been diagnosed. Unlike most proposed dementia interventions, Shingrix is already licensed, relatively inexpensive compared to approved alternatives, and routinely used in older adults. Our findings from a large matched real-world cohort, together with supportive concordant analyses of repeated MMSE measurements, raise the possibility that recombinant zoster vaccination could represent a practical and underexplored strategy for improving outcomes in patients with established dementia. Given the limited therapeutic options currently available, the scale of the public health burden, and the feasibility of implementation in routine care, these findings justify prospective clinical testing. Importantly, the commercial incentive to pursue definitive trials in this setting may still be modest relative to the potential societal benefit, because this would involve evaluating an already available vaccine in a new clinical context rather than developing an entirely new dementia therapeutic. That imbalance between possible public-health impact and commercial return should not diminish the importance of rigorously evaluating this prospective approach.

## Online Methods

### Study population and cohort assembly

We conducted a retrospective observational cohort study using de-identified electronic health record data from the TriNetX US Collaborative Network. The analyses were carried out from February 2026 to March 2026 on data available from 114,295,757 individuals including 66 healthcare organizations (HCOs). The source population comprised adults with a recorded diagnosis of dementia. Dementia was defined broadly and included Alzheimer’s disease (G30.0), vascular dementia (F01), frontotemporal dementia (G31.0), dementia with Lewy bodies (G31.83), dementia in other diseases classified elsewhere (F02) and unspecified dementia (F03), encompassing the major neurodegenerative disorders captured by routine coding in the electronic health record (EHR) network. Dementia diagnoses were identified using ICD codes, which have been previously validated in administrative and electronic health record data, demonstrating high specificity and moderate sensitivity^34,35^. The initial query yielded 1,487,167 patients with one or more qualifying dementia codes. Of these, 34,543 met criteria for the Shingrix-exposed cohort, and 200,176 met criteria for the active comparator cohort vaccinated by a non-Shingrix vaccine. In the unvaccinated cohort 941,374 individuals were included before matching. Cohorts included all individuals who received Shingrix vaccine or any other vaccine between 20 October 2017 and 31 December 2025 or visited the healthcare provider during the same period of time and did not have an encounter for immunization. The date of the first dementia diagnosis code in EHR was considered the index date.

### Exposure, comparator and follow-up definitions

Exposure was defined as receipt of recombinant zoster vaccine Shingrix within two years of a recorded dementia diagnosis. Shingrix was identified in EHR using the RxNorm code 1986821 and the first recorded dose was used to satisfy the exposure criteria. The index event remained the date of first dementia diagnosis in all groups, and a 5 year follow-up began only after the 2-year vaccination window to avoid immortal time bias,.

The comparator cohort consisted of patients with dementia who received other vaccines during the same two-year post-diagnosis window defined as individuals encountering immunization (ICD-10 code Z23), but had no record of Shingrix during that ascertainment period. This ICD-10 code encompassed individuals receiving any of the following vaccines: Flu (CVX 88, 123, 128, RxNorm 1303851, 857917, 857919, 1005929), Tdap (CVX 115), Pneumopcv (CVX 152), Sars-cov-2 (covid- 19) (CVX 213), Hepb (CVX 45), Td (CVX 139), Dtap (CVX107), Polio (CvX 89), Hib (CVX 17), Pneumoppv (CVX 33), Mmr (CVX 3), Varicella (CVX 21), Streptococcus pneumoniae (RxNorm 2566291 – 2566297, 2686917-2686927), Hepa (CVX 85), RSV (CVX 304, RxNorm 2636589, 2685002), Bcg (CVX 19), Rabies (CVX 90), Mening (CVX 108), Hepatitis (RxNorm 253174), Hpv (CVX 137), Yellowfever (CVX 37), Rotavirus (CVX 122), Adeno (CVX 82), and the live zoster vaccine Zostavax (RxNorm 1292422 or CVX 121), however the group is defined in the period in which the new recombinant vaccine Shingrix became available and Zostavax discontinued shortly after. The broad intent of the comparator was to restrict the analysis to vaccinated individuals, thereby reducing bias from differential healthcare contact or willingness to accept preventive care.

### Covariates

Cohorts were matched for 43 covariates spanning all sociodemographic characteristics, comorbidity burden, lifestyle proxies, healthcare-utilization proxies and socioeconomic indicators recorded any time prior to the index date up to and including the index date.

These variables included age at index event, sex, race, ethnicity and marital status; body mass index (BMI; LOINC 9083) and blood pressure categories (LOINC 9085, 9086); ICD-10 codes for smoking (Z72.0, Z87.89) and alcohol abuse (F10.1). Cohorts were balanced across major ICD-10 categories including diabetes mellitus (E08-E13), Parkinson’s disease (G20), ischemic heart diseases (I20-I25), other forms of heart disease (deprecated 2021) (I30-I52), heart failure (I50), cerebrovascular diseases (I60-I69), other chronic obstructive pulmonary disease (J44), unspecified cirrhosis of liver (K74.60) and chronic kidney disease (N18).

Matching also incorporated proxies for healthcare utilization and socioeconomic context, including routine medical examination codes (Z00.00, Z00.01), cancer screening (Z12), long-term drug therapy (Z79), counseling for socioeconomic factors (Z56) and State Medicaid (T codes) and private payer (S codes), and problems related to housing and economic circumstances (Z59). Covariate balance was assessed using standardized mean differences (SMD).

Laboratory measurements were available as continuous variables summarized by their mean values; however, to improve matching precision and to avoid imputation of missing values, these variables were categorized into clinically relevant bins. BMI was grouped as ≤18.4, 18.5–24.9, 25–29.9 and ≥30 kg m⁻²; systolic blood pressure as ≤119, 120–129, 130–139, 140–179 and >180 mmHg; and diastolic blood pressure as ≤79, 80–89, 90–119 and ≥120 mmHg. For MMSE (LOINC 72106-8) the categories were 0-9, 10-20, 21-23, and 24-30. Missing categorical covariates are retained as an unknown category. No specific imputation was implemented for missing data. In the Cox regression (model 6), each covariate HR compares participants with the indicated code or category with all participants without that code or category.

### Outcomes

The primary outcome was all-cause mortality captured within the EHR network. Because cause-of-death information is often incomplete in real-world EHR research, and unavailable in TriNetX, all-cause mortality was used.

Secondary negative control outcomes to assess specificity of the observed association included asthma (J45), cerebral infarction (I63), fractures of wrist (S62) or femur (S72) and admission to nursing home (Z59.3).

### Statistical analysis

We followed patients starting two years after the time of dementia diagnosis for 5 years, until an outcome occurred, the end of the study period, or the last recorded activity in the database, whichever occurred first. Descriptive statistics were used to summarize baseline sociodemographic and clinical characteristics of each cohort. Propensity score matching (PSM) was used through the TriNetX built-in algorithm that includes logistic regression implemented by the function Logistic Regression of the scikit-learn package in Python version 3.7 (Python Software Foundation). All listed covariates were used for PSM 1:1, based on a greedy nearest neighbor. The matching itself was performed with numpy 1.21.5 in Python 3.7. with a caliper of 0.1. A two-sided p-value ≤0.05 was considered statistically significant; characteristics with the SMD<0.1 after matching were considered well-matched^36^. To avoid potential bias from covariates ordering, the records were randomized before matching.

Outcomes were compared between the PSM cohorts over time using Kaplan-Meier analysis with the log-rank test calculated using the R survival package version 3.2.3 (R Foundation for Statistical Computing). Hazard ratios (HRs) were calculated using a proportional hazard model wherein the cohort to which the patient belonged was used as the independent variable. HRs were assessed for proportionality using the generalized Schoenfeld approach where a P ≤0.05 was considered significant for nonproportionality.

### Repeated MMSE analyses

To explore cognitive trajectory, we used TriNetX cohort definitions to identify patients with two recorded MMSE measurements 3 to 6 years apart. Cohorts were stratified according to vaccination status in the interval between the two MMSE measurements: no recorded vaccination, Shingrix vaccination, or vaccination with non-Shingrix vaccines, all after 20 October 2017 as in our mortality analysis. For vaccinated cohorts, the pre-vaccination MMSE was defined as the first qualifying MMSE before vaccination and the post-vaccination MMSE as the most recent qualifying MMSE after vaccination. Cohort logic required that vaccination occurred between the two MMSE assessments and that the two assessments were separated by 3 to 6 years. For unvaccinated controls, analogous first and last MMSE measurements 3 to 6 years apart were identified with any type of visit to the healthcare provider recorded between the two MMSE measurements but no recorded vaccination throughout the period.

### Ethics statement

All TriNetX data are deidentified and anonymized in compliance with the Health Insurance Portability and Accountability Act; thus, informed consent was not necessary, and the study was given an exemption from the Hadassah Medical Center Institutional Review Board.

## Data Availability

The data used in this study are available through the TriNetX platform to institutions with a licensed agreement. Due to patient privacy and data sharing agreements, individual-level data cannot be shared directly by the authors.

## Code Availability

Data visualization were performed using R version 3.2.3. All custom scripts utilized to generate the figures presented in this manuscript, including the violin plots, forest plots, and survival plots, will be made publicly available via a dedicated public repository upon manuscript acceptance.

## Author contributions

MSBH, CLG, RCM, BYK, JS, and KS designed the study and analyses; KS, RSB, and MSBH performed analyses; MSBH wrote the manuscript with extensive feedback and editing from NIS, KS, JS, BYK, RCM, and CLG.

---

**Table S1.** Baseline characteristics of the main dementia cohort before and after propensity score matching. Characteristics are shown for patients vaccinated with Shingrix and comparator patients vaccinated with non-Shingrix vaccines. The percentage of individuals from the entire cohort is in prentices (%n). Balance is summarized using standardized mean differences (SMDs).

|  | Before matching |  |  | After matching |  |  |
| --- | --- | --- | --- | --- | --- | --- |
|  | Shingrix<br>vaccinated<br>34,573 | Non-Shingrix<br>vaccinated<br>197,581 | SMD | Shingrix<br>vaccinated<br>34,480 | Non-Shingrix<br>vaccinated<br>34,480 | SMD |
| **Number of participants** |  |  |  |  |  |  |
| **SOCIODEMOGRAPHICS** |  |  |  |  |  |  |
| Age at Index (AI) | 75.4 (9.5) | 77.5 (10.8) | 0.202 | 75.4 (9.5) | 75 (12.9) | 0.041 |
| **Sex; n (%)** |  |  |  |  |  |  |
| Female (F) | 20278 (58.8) | 119630 (60.6) | 0.037 | 20278 (58.8) | 20360 (59) | 0.005 |
| Male (M) | 14186 (41.1) | 77741 (39.4) | 0.036 | 14186 (41.1) | 14101 (40.9) | 0.005 |
| Unknown Gender (UN) | 16 (0.1) | 43 (0.02) | 0.013 | 16 (0.046) | 19 (0.1) | 0.004 |
| **Race; n (%)** |  |  |  |  |  |  |
| White (2106-3) | 24364 (70.7) | 150199 (76.1) | 0.123 | 24364 (70.7) | 24177 (70.1) | 0.012 |
| Black or African American (2054-5) | 4832 (14.0) | 26583 (13.5) | 0.016 | 4832 (14) | 4853 (14.1) | 0.002 |
| Asian (2028-9) | 2883 (8.4) | 9149 (4.6) | 0.152 | 2883 (8.4) | 3044 (8.8) | 0.017 |
| Native Hawaiian or Other Pacific Islander (2076-8) | 387 (1.1) | 1026 (0.5) | 0.067 | 387 (1.1) | 425 (1.2) | 0.01 |
| American Indian or Alaska Native (1002-5) | 109 (0.3) | 475 (0.2) | 0.014 | 109 (0.3) | 102 (0.3) | 0.004 |
| Other Race (2131-1) | 869 (2.5) | 4333 (2.2) | 0.021 | 869 (2.5) | 908 (2.6) | 0.007 |
| Unknown Race (UNK) | 1036 (3.0) | 5649 (2.9) | 0.009 | 1036 (3) | 971 (2.8) | 0.011 |
| **Ethnicity; n (%)** |  |  |  |  |  |  |
| Hispanic or Latino (2135-2) | 1921 (5.6) | 7279 (3.7) | 0.09 | 1921 (5.6) | 2027 (5.9) | 0.013 |
| Not Hispanic or Latino (2186-5) | 29446 (85.4) | 142837 (72.4) | 0.324 | 29446 (85.4) | 29032 (84.2) | 0.033 |
| **Marital status; n (%)** |  |  |  |  |  |  |
| Married (M) | 9489 (27.5) | 56021 (28.4) | 0.019 | 9489 (27.5) | 9435 (27.4) | 0.004 |
| Never Married (S) | 2992 (8.7) | 18932 (9.6) | 0.032 | 2992 (8.7) | 3050 (8.8) | 0.006 |
| Divorced (D) | 1921 (5.6) | 12318 (6.2) | 0.028 | 1921 (5.6) | 1904 (5.5) | 0.002 |
| Widowed (W) | 6174 (17.9) | 47408 (24.0) | 0.151 | 6174 (17.9) | 6144 (17.8) | 0.002 |
| Domestic partner (T) | 136 (0.4) | 491 (0.3) | 0.026 | 136 (0.4) | 133 (0.4) | 0.001 |
| Legally Separated (L) | 168 (0.5) | 1062 (0.5) | 0.007 | 168 (0.5) | 183 (0.5) | 0.006 |
| **Lifestyle; n (%)** |  |  |  |  |  |  |
| Tobacco use (Z72.0) | 1574 (4.6) | 10158 (5.2) | 0.027 | 1574 (4.6) | 1591 (4.6) | 0.002 |
| Personal history of nicotine dependence (Z87.891) | 7669 (22.2) | 51028 (25.9) | 0.085 | 7669 (22.2) | 7466 (21.7) | 0.014 |
| Alcohol abuse (F10.1) | 1461 (4.2) | 8073 (4.1) | 0.007 | 1461 (4.2) | 1405 (4.1) | 0.008 |
| **Health-seeking behavior; n (%)** |  |  |  |  |  |  |
| Encounter for general adult medical examination without abnormal findings (Z00.00) | 13995 (40.6) | 88375 (44.8) | 0.085 | 13995 (40.6) | 14592 (42.3) | 0.035 |
| Encounter for general adult medical examination with abnormal findings (Z00.01) | 921 (2.7) | 6910 (3.5) | 0.048 | 921 (2.7) | 897 (2.6) | 0.004 |
| Long term (current) drug therapy (Z79) | 14925 (43.3) | 112765 (57.1) | 0.279 | 14925 (43.3) | 14712 (42.7) | 0.013 |
| Encounter for screening for malignant neoplasms (Z12) | 13554 (39.3) | 79296 (40.2) | 0.018 | 13554 (39.3) | 13966 (40.5) | 0.024 |
| **Social determinants of health; n (%)** |  |  |  |  |  |  |
| Problems related to employment and unemployment (Z56) | 122 (0.4) | 786 (0.4) | 0.007 | 122 (0.4) | 120 (0.3) | 0.001 |
| Problems related to housing and economic circumstances (Z59) | 570 (1.7) | 4441 (2.3) | 0.043 | 570 (1.7) | 585 (1.7) | 0.003 |
| S: Private Payer Codes | 776 (2.3) | 5367 (2.7) | 0.03 | 776 (2.3) | 845 (2.5) | 0.013 |
| T: State Medicaid Agency Codes | 80 (0.2) | 477 (0.2) | 0.002 | 80 (0.2) | 85 (0.2) | 0.003 |
| **COMORBIDITIES [ICD-10 code]; n (%)** |  |  |  |  |  |  |
| Diabetes mellitus (E08-E13) | 11418 (33.1) | 67214 (34.1) | 0.02 | 11418 (33.1) | 11226 (32.6) | 0.012 |
| Parkinson's disease (G20) | 1834 (5.3) | 11446 (5.8) | 0.021 | 1834 (5.3) | 1780 (5.2) | 0.007 |
| Ischemic heart diseases (I20-I25) | 9761 (28.3) | 64803 (32.8) | 0.098 | 9761 (28.3) | 9487 (27.5) | 0.018 |
| Other forms of heart disease (deprecated 2021) (I30-I52) | 15386 (44.6) | 99842 (50.6) | 0.119 | 15386 (44.6) | 15177 (44) | 0.012 |
| Heart failure (I50) | 4861 (14.1) | 38269 (19.4) | 0.142 | 4861 (14.1) | 4749 (13.8) | 0.009 |
| Cerebrovascular diseases (I60-I69) | 9607 (27.9) | 60493 (30.6) | 0.061 | 9607 (27.9) | 9447 (27.4) | 0.01 |
| Other chronic obstructive pulmonary disease (J44) | 4536 (13.2) | 31259 (15.8) | 0.076 | 4536 (13.2) | 4481 (13) | 0.005 |
| Unspecified cirrhosis of liver (K74.60) | 491 (1.4) | 3308 (1.7) | 0.02 | 491 (1.4) | 462 (1.3) | 0.007 |
| Chronic kidney disease (N18) | 7304 (21.2) | 49093 (24.9) | 0.088 | 7304 (21.2) | 7023 (20.4) | 0.02 |

| **Labs** |  |  |  |  |  |  |
| --- | --- | --- | --- | --- | --- | --- |
| *BMI (9083)* | 27195 (78.9) | 142917 (72.4) |  | 27195 (78.9) | 26456 (76.7) |  |
| At most 18.4 kg/m2 | 2456 (7.1) | 14592 (7.4) | 0.01 | 2456 (7.1) | 2480 (7.2) | 0.003 |
| 18.5-24.9 kg/m2 | 13722 (39.8) | 76524 (38.8) | 0.021 | 13722 (39.8) | 13646 (39.6) | 0.005 |
| 25-29.9 kg/m2 | 16240 (47.1) | 86178 (43.7) | 0.069 | 16240 (47.1) | 16162 (46.9) | 0.005 |
| ≥30 kg/m2 | 12017 (34.9) | 63029 (31.9) | 0.062 | 12017 (34.9) | 11899 (34.5) | 0.007 |
| *Blood Pressure, Systolic (9085)* | 29927 (86.8) | 161728 (81.9) |  | 29927 (86.8) | 29876 (86.6) |  |
| At most 119 mm[Hg] | 22749 (66.0) | 121751 (61.7) | 0.09 | 22749 (66) | 22648 (65.7) | 0.006 |
| 120 - 129 mm[Hg] | 23861 (69.2) | 126327 (63.99) | 0.111 | 23861 (69.2) | 23817 (69.1) | 0.003 |
| 130 - 139 mm[Hg] | 23857 (69.2) | 126441 (64.1) | 0.109 | 23857 (69.2) | 23716 (68.8) | 0.009 |
| 140 - 179 mm[Hg] | 24275 (70.4) | 130577 (66.1) | 0.092 | 24275 (70.4) | 24144 (70) | 0.008 |
| ≥180 mm[Hg] | 7812 (22.7) | 46563 (23.6) | 0.022 | 7812 (22.7) | 7746 (22.5) | 0.005 |
| *Blood Pressure, Diastolic (9086)* | 29930 (86.8) | 161725 (81.9) |  | 29930 (86.8) | 29876 (86.6) |  |
| At most 79 mm[Hg] | 28314 (82.1) | 152944 (77.5) | 0.116 | 28314 (82.1) | 28177 (81.7) | 0.01 |
| 80 - 89 mm[Hg] | 23529 (68.2) | 124196 (62.9) | 0.112 | 23529 (68.2) | 23410 (67.9) | 0.007 |
| 90 - 119 mm[Hg] | 15478 (44.9) | 83498 (42.3) | 0.052 | 15478 (44.9) | 15356 (44.5) | 0.007 |
| ≥120 mm[Hg] | 1235 (3.6) | 7556 (3.8) | 0.013 | 1235 (3.6) | 1222 (3.5) | 0.002 |
| *Total score [MMSE] (72106-8)* | 31 (0.09) | 423 (0.2) |  | 31 (0.1) | 32 (0.1) |  |
| At most 9 [score] | 10 (0.03) | 11 (0.01) | 0.018 | 10 (0.03) | 10 (0.03) | < 0.0001 |
| 10-20 [score] | 10 (0.03) | 125 (0.06) | 0.016 | 10 (0.03) | 13 (0.04) | 0.005 |
| 21-23 [score] | 10 (0.03) | 91 (0.05) | 0.009 | 10 (0.03) | 11 (0.03) | 0.002 |
| 24-30 [score] | 21 (0.06) | 259 (0.1) | 0.023 | 21 (0.1) | 15 (0.04) | 0.008 |

**Table S2.** Baseline characteristics of the high socioeconomic status subgroup before and after propensity score matching. Characteristics are shown for patients vaccinated with Shingrix and comparator patients vaccinated with non-Shingrix vaccines in the subgroup defined by a recorded cosmetic surgery as a proxy for higher socioeconomic status. The percentage of individuals from the entire cohort is in prentices (%n), and balance is summarized using standardized mean differences (SMDs).

| Number of participants | Before matching |  |  | After matching |  |  |
| --- | --- | --- | --- | --- | --- | --- |
|  | Shingrix<br>vaccinated<br>1033 | Non-Shingrix<br>vaccinated<br>3892 | SMD | Shingrix<br>vaccinated<br>1001 | Non-Shingrix<br>vaccinated<br>1001 | SMD |
| **SOCIODEMOGRAPHICS** |  |  |  |  |  |  |
| Age at Index (AI) | 71 (10.5) | 70.4 (15.8) | 0.04 | 70.9 (10.5) | 71.2 (14.6) | 0.02 |
| **Sex; n (%)** |  |  |  |  |  |  |
| Female (F) | 658 (64) | 2483 (63.8) | 0.004 | 646 (64.5) | 657 (65.6) | 0.02 |
| Male (M) | 370 (36) | 1407 (36.2) | 0.004 | 355 (35.5) | 344 (34.4) | 0.02 |
| Unknown Gender (UN) | 0 (0) | 0 (0) | - | 0 (0) | 0 (0) | - |
| **Race; n (%)** |  |  |  |  |  |  |
| White (2106-3) | 772 (75.1) | 3012 (77.4) | 0.06 | 760 (75.9) | 749 (74.8) | 0.03 |
| Black or African American (2054-5) | 125 (12.2) | 423 (10.9) | 0.04 | 117 (11.7) | 135 (13.5) | 0.05 |
| Asian (2028-9) | 60 (5.8) | 178 (4.6) | 0.06 | 58 (5.8) | 58 (5.8) | < 0.0001 |
| Native Hawaiian or Other Pacific Islander (2076-8) | 10 (1) | 13 (0.3) | 0.08 | 10 (1) | 10 (1) | < 0.0001 |
| American Indian or Alaska Native (1002-5) | 10 (1) | 13 (0.3) | 0.08 | 10 (1) | 10 (1) | < 0.0001 |
| Other Race (2131-1) | 36 (3.5) | 123 (3.2) | 0.02 | 32 (3.2) | 29 (2.9) | 0.02 |
| Unknown Race (UNK) | 29 (2.8) | 128 (3.3) | 0.03 | 29 (2.9) | 26 (2.6) | 0.02 |
| **Ethnicity; n (%)** |  |  |  |  |  |  |
| Hispanic or Latino (2135-2) | 73 (7.1) | 181 (4.7) | 0.1 | 61 (6.1) | 63 (6.3) | 0.01 |
| Not Hispanic or Latino (2186-5) | 861 (83.8) | 3084 (79.3) | 0.12 | 846 (84.5) | 819 (81.8) | 0.07 |
| **Marital status; n (%)** |  |  |  |  |  |  |
| Married (M) | 274 (26.7) | 1250 (32.1) | 0.12 | 273 (27.3) | 276 (27.6) | 0.01 |
| Never Married (S) | 80 (7.8) | 454 (11.7) | 0.13 | 80 (8) | 80 (8) | < 0.0001 |
| Divorced (D) | 74 (7.2) | 270 (6.9) | 0.01 | 74 (7.4) | 80 (8) | 0.02 |
| Widowed (W) | 145 (14.1) | 667 (17.1) | 0.08 | 144 (14.4) | 146 (14.6) | 0.01 |
| Domestic partner (T) | 10 (1) | 18 (0.5) | 0.06 | 10 (1) | 10 (1) | < 0.0001 |
| Legally Separated (L) | 10 (1) | 19 (0.5) | 0.06 | 10 (1) | 10 (1) | < 0.0001 |
| **Lifestyle; n (%)** |  |  |  |  |  |  |
| Tobacco use (Z72.0) | 67 (6.5) | 283 (7.3) | 0.03 | 65 (6.5) | 68 (6.8) | 0.01 |
| Personal history of nicotine dependence (Z87.891) | 303 (29.5) | 1289 (33.1) | 0.08 | 300 (30) | 302 (30.2) | 0.004 |
| Alcohol abuse (F10.1) | 56 (5.4) | 211 (5.4) | 0.001 | 49 (4.9) | 56 (5.6) | 0.03 |
| **Health-seeking behavior; n (%)** |  |  |  |  |  |  |
| Encounter for general adult medical examination without abnormal findings (Z00.00) | 511 (49.7) | 2220 (57.1) | 0.15 | 505 (50.5) | 507 (50.6) | 0.004 |
| Encounter for general adult medical examination with abnormal findings (Z00.01) | 42 (4.1) | 252 (6.5) | 0.11 | 42 (4.2) | 39 (3.9) | 0.02 |
| Long term (current) drug therapy (Z79) | 595 (57.9) | 2830 (72.8) | 0.32 | 595 (59.4) | 582 (58.1) | 0.03 |
| Encounter for screening for malignant neoplasms (Z12) | 543 (52.8) | 2252 (57.9) | 0.1 | 537 (53.6) | 549 (54.8) | 0.02 |
| **Social determinants of health; n (%)** |  |  |  |  |  |  |
| Problems related to employment and unemployment (Z56) | 10 (1) | 43 (1.1) | 0.01 | 10 (1) | 10 (1) | < 0.0001 |
| Problems related to housing and economic circumstances (Z59) | 18 (1.8) | 137 (3.5) | 0.11 | 18 (1.8) | 30 (3) | 0.08 |
| S: Private Payer Codes | 51 (5) | 312 (8) | 0.12 | 51 (5.1) | 46 (4.6) | 0.02 |
| T: State Medicaid Agency Codes | 10 (1) | 25 (0.6) | 0.04 | 10 (1) | 10 (1) | < 0.0001 |
| **COMORBIDITIES [ICD-10 code]; n (%)** |  |  |  |  |  |  |
| Diabetes mellitus (E08-E13) | 362 (35.2) | 1387 (35.7) | 0.01 | 349 (34.9) | 365 (36.5) | 0.03 |
| Parkinson's disease (G20) | 151 (14.7) | 583 (15) | 0.01 | 151 (15.1) | 134 (13.4) | 0.05 |
| Ischemic heart diseases (I20-I25) | 324 (31.5) | 1384 (35.6) | 0.09 | 318 (31.8) | 311 (31.1) | 0.02 |
| Other forms of heart disease (deprecated 2021) (I30-I52) | 507 (49.3) | 2253 (57.9) | 0.17 | 498 (49.8) | 487 (48.7) | 0.02 |
| Heart failure (I50) | 147 (14.3) | 814 (20.9) | 0.18 | 144 (14.4) | 152 (15.2) | 0.02 |
| Cerebrovascular diseases (I60-I69) | 385 (37.5) | 1597 (41.1) | 0.07 | 377 (37.7) | 389 (38.9) | 0.03 |
| Other chronic obstructive pulmonary disease (J44) | 145 (14.1) | 661 (17) | 0.08 | 145 (14.5) | 145 (14.5) | < 0.0001 |
| Unspecified cirrhosis of liver (K74.60) | 18 (1.8) | 86 (2.2) | 0.03 | 18 (1.8) | 18 (1.8) | < 0.0001 |
| Chronic kidney disease (N18) | 218 (21.2) | 1001 (25.7) | 0.11 | 213 (21.3) | 218 (21.8) | 0.01 |
| **Labs** |  |  |  |  |  |  |
| *BMI (9083)* | 854 (83.1) | 3009 (77.4) |  | 829 (82.8) | 793 (79.2) |  |
| At most 18.4 kg/m2 | 102 (9.9) | 430 (11.1) | 0.04 | 97 (9.7) | 107 (10.7) | 0.03 |
| 18.5-24.9 kg/m2 | 439 (42.7) | 1706 (43.9) | 0.02 | 430 (43) | 421 (42.1) | 0.02 |
| 25-29.9 kg/m2 | 526 (51.2) | 1984 (51) | 0.003 | 508 (50.7) | 517 (51.6) | 0.02 |
| ≥ 30 kg/m2 | 459 (44.7) | 1653 (42.5) | 0.04 | 442 (44.2) | 428 (42.8) | 0.03 |
| *Blood Pressure, Systolic (9085)* | 916 (89.1) | 3367 (86.6) |  | 889 (88.8) | 893 (89.2) |  |
| At most 119 mm[Hg] | 811 (78.9) | 3043 (78.2) | 0.02 | 785 (78.4) | 792 (79.1) | 0.02 |
| 120 - 129 mm[Hg] | 814 (79.2) | 3058 (78.6) | 0.01 | 787 (78.6) | 786 (78.5) | 0.002 |
| 130 - 139 mm[Hg] | 791 (76.9) | 2998 (77.1) | 0.003 | 768 (76.7) | 773 (77.2) | 0.01 |
| 140 - 179 mm[Hg] | 779 (75.8) | 2940 (75.6) | 0.01 | 755 (75.4) | 758 (75.7) | 0.01 |
| ≥180 mm[Hg] | 264 (25.7) | 1129 (29) | 0.08 | 261 (26.1) | 259 (25.9) | 0.01 |
| *Blood Pressure, Diastolic (9086)* | 916 (89.1) | 3367 (86.6) |  | 889 (88.8) | 893 (89.2) |  |
| At most 79 mm[Hg] | 901 (87.6) | 3305 (85) | 0.08 | 874 (87.3) | 878 (87.7) | 0.01 |
| 80 - 89 mm[Hg] | 781 (76) | 3009 (77.4) | 0.03 | 759 (75.8) | 754 (75.3) | 0.01 |
| 90 - 119 mm[Hg] | 596 (58) | 2301 (59.2) | 0.02 | 580 (57.9) | 588 (58.7) | 0.02 |
| ≥120 mm[Hg] | 46 (4.5) | 293 (7.5) | 0.13 | 46 (4.6) | 53 (5.3) | 0.03 |
| *Total score [MMSE] (72106-8)* | 10 (1) | 10 (0.3) |  | 10 (1) | 0 (0) |  |
| At most 9 [score] | 0 (0) | 0 (0) | - | 0 (0) | 0 (0) | - |
| 10-20 [score] | 0 (0) | 10 (0.3) | 0.07 | 0 (0) | 0 (0) | - |
| 21-23 [score] | 0 (0) | 0 (0) | - | 0 (0) | 0 (0) | - |
| 24-30 [score] | 10 (1) | 10 (0.3) | 0.09 | 10 (1) | 0 (0) | 0.14 |

**Table S3.** Cox proportional hazards model (Model 6) for all-cause mortality in the un-matched cohorts. Hazard ratios (HRs), 95% confidence intervals (CIs), and P values are shown for the association of vaccination status (Shingrix-vaccinated vs. Non-Shingrix vaccinated) and covariates with all-cause mortality in model6 (related to Fig.3). Each HR compares participants with the indicated code or category with all participants without that code or category.

|  | Hazard Ratio | 95% CI |  | P > \|z\| |
| --- | --- | --- | --- | --- |
|  |  | Lower | Higher |  |
| Vaccination group (Shingrix vs non-Shingrix) | 0.81 | 0.788 | 0.832 | < 0.0001 |
| Male (M) | 1.282 | 1.255 | 1.311 | < 0.0001 |
| Age at Index (AI) | 1.055 | 1.054 | 1.057 | < 0.0001 |
| White (2106-3) | 1.247 | 1.159 | 1.342 | < 0.0001 |
| Black or African American (2054-5) | 0.865 | 0.8 | 0.936 | 0.0003 |
| Asian (2028-9) | 0.849 | 0.78 | 0.925 | 0.0002 |
| American Indian or Alaska Native (1002-5) | 1.124 | 0.891 | 1.418 | 0.3222 |
| Native Hawaiian or Other Pacific Islander (2076-8) | 1.063 | 0.924 | 1.222 | 0.3916 |
| Other Race (2131-1) | 0.919 | 0.824 | 1.024 | 0.1272 |
| *BMI (9083)* |  |  |  |  |
| At most 18.4 kg/m2 | 1.264 | 1.214 | 1.317 | < 0.0001 |
| 18.5-24.9 kg/m2 | 1.106 | 1.078 | 1.135 | < 0.0001 |
| 25-29.9 kg/m2 | 0.988 | 0.965 | 1.012 | 0.3339 |
| ≥30 kg/m2 | 1.014 | 0.986 | 1.043 | 0.3351 |
| Tobacco use (Z72.0) | 1.255 | 1.191 | 1.323 | < 0.0001 |
| Personal history of nicotine dependence (Z87.891) | 1.068 | 1.04 | 1.096 | < 0.0001 |
| Alcohol abuse (F10.1) | 0.979 | 0.922 | 1.038 | 0.4721 |
| Encounter for general adult medical examination without abnormal findings (Z00.00) | 1.045 | 1.022 | 1.07 | 0.0001 |
| Encounter for general adult medical examination with abnormal findings (Z00.01) | 0.965 | 0.898 | 1.037 | 0.3279 |
| Long term (current) drug therapy (Z79) | 1.066 | 1.041 | 1.092 | < 0.0001 |
| Encounter for screening for malignant neoplasms (Z12) | 0.911 | 0.889 | 0.933 | < 0.0001 |
| Problems related to employment and unemployment (Z56) | 0.896 | 0.691 | 1.162 | 0.4084 |
| Problems related to housing and economic circumstances (Z59) | 1.154 | 1.046 | 1.274 | 0.0044 |
| T: State Medicaid Agency Codes (T) | 0.78 | 0.577 | 1.053 | 0.1046 |
| S: Private Payer Codes (S) | 1.031 | 0.961 | 1.106 | 0.3955 |
| *Blood Pressure, Systolic (9085)* |  |  |  |  |
| At most 119 mm[Hg] | 1.054 | 1.021 | 1.088 | 0.0013 |
| 120 - 129 mm[Hg] | 0.986 | 0.952 | 1.021 | 0.4157 |
| 130 - 139 mm[Hg] | 0.955 | 0.922 | 0.99 | 0.0112 |
| 140 - 179 mm[Hg] | 0.909 | 0.878 | 0.942 | < 0.0001 |
| ≥180 mm[Hg] | 1.037 | 1.008 | 1.067 | 0.0125 |
| *Blood Pressure, Diastolic (9086)* |  |  |  |  |
| At most 79 mm[Hg] | 0.979 | 0.939 | 1.021 | 0.3251 |
| 80 - 89 mm[Hg] | 0.987 | 0.954 | 1.022 | 0.4695 |
| 90 - 119 mm[Hg] | 1.018 | 0.99 | 1.047 | 0.2019 |
| ≥120 mm[Hg] | 1.065 | 1.002 | 1.132 | 0.0444 |
| Diabetes mellitus (E08-E13) | 1.088 | 1.064 | 1.114 | < 0.0001 |
| Cerebrovascular diseases (I60-I69) | 1.012 | 0.989 | 1.036 | 0.3065 |
| Ischemic heart diseases (I20-I25) | 1.039 | 1.014 | 1.064 | 0.0021 |
| Other forms of heart disease (deprecated 2021) (I30-I52) | 1.148 | 1.12 | 1.177 | < 0.0001 |
| Heart failure (I50) | 1.198 | 1.163 | 1.235 | < 0.0001 |
| Chronic kidney disease (N18) | 1.291 | 1.259 | 1.323 | < 0.0001 |
| Unspecified cirrhosis of liver (K74.60) | 1.355 | 1.242 | 1.479 | < 0.0001 |
| Other chronic obstructive pulmonary disease (J44) | 1.153 | 1.12 | 1.187 | < 0.0001 |
| Parkinson's disease (G20) | 1.398 | 1.341 | 1.457 | < 0.0001 |
| *Total score [MMSE] (72106-8) \* MMSE scores exist for less than 0.2% of the cohort* |  |  |  |  |
| At most 9 [score] | - |  |  | - |
| 10-20 [score] | 0.269 | 0.101 | 0.716 | 0.0086 |
| 21-23 [score] | 0.97 | 0.403 | 2.333 | 0.9455 |
| 24-30 [score] | 0.303 | 0.144 | 0.636 | 0.0016 |

**Table S4.**
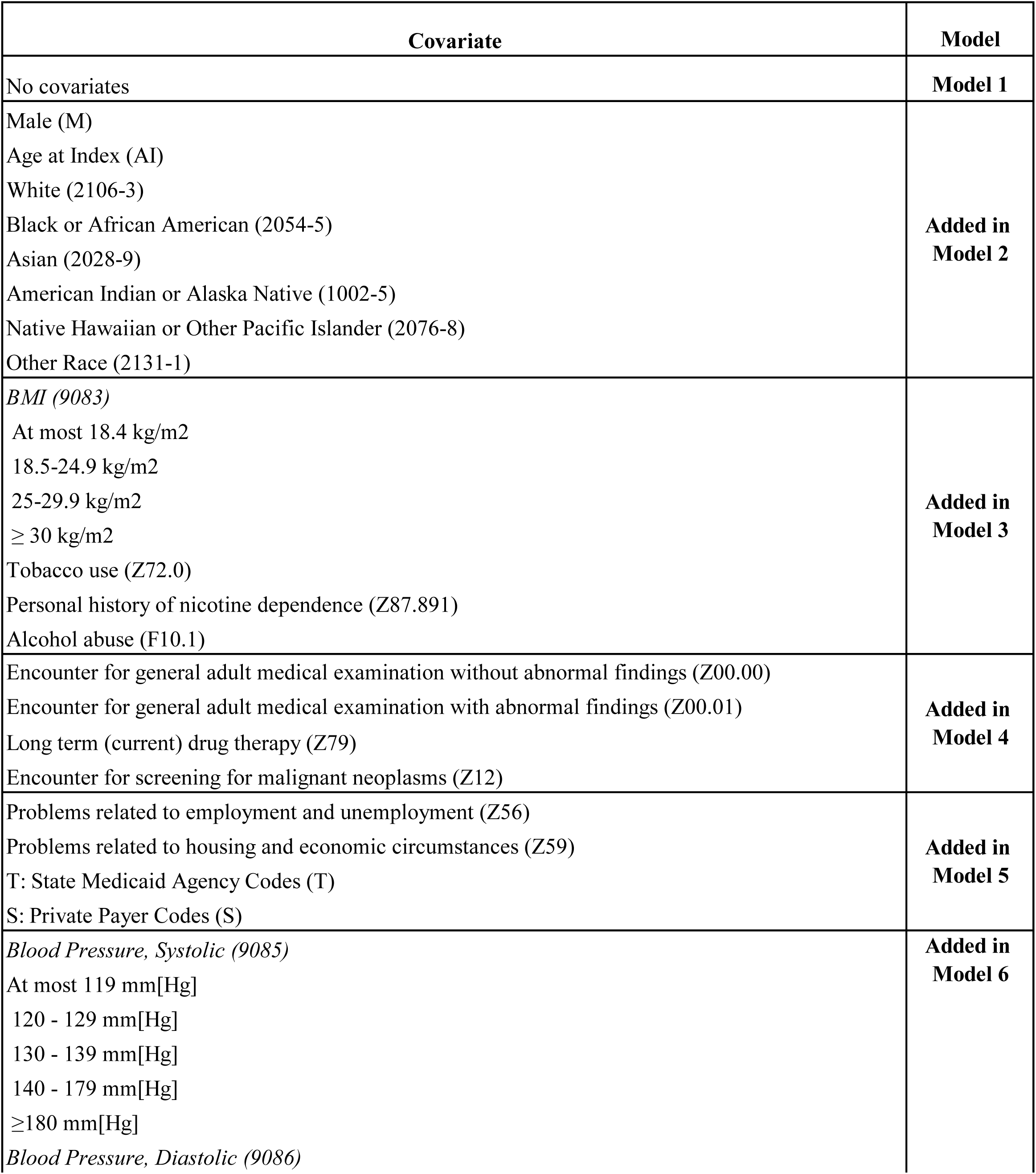
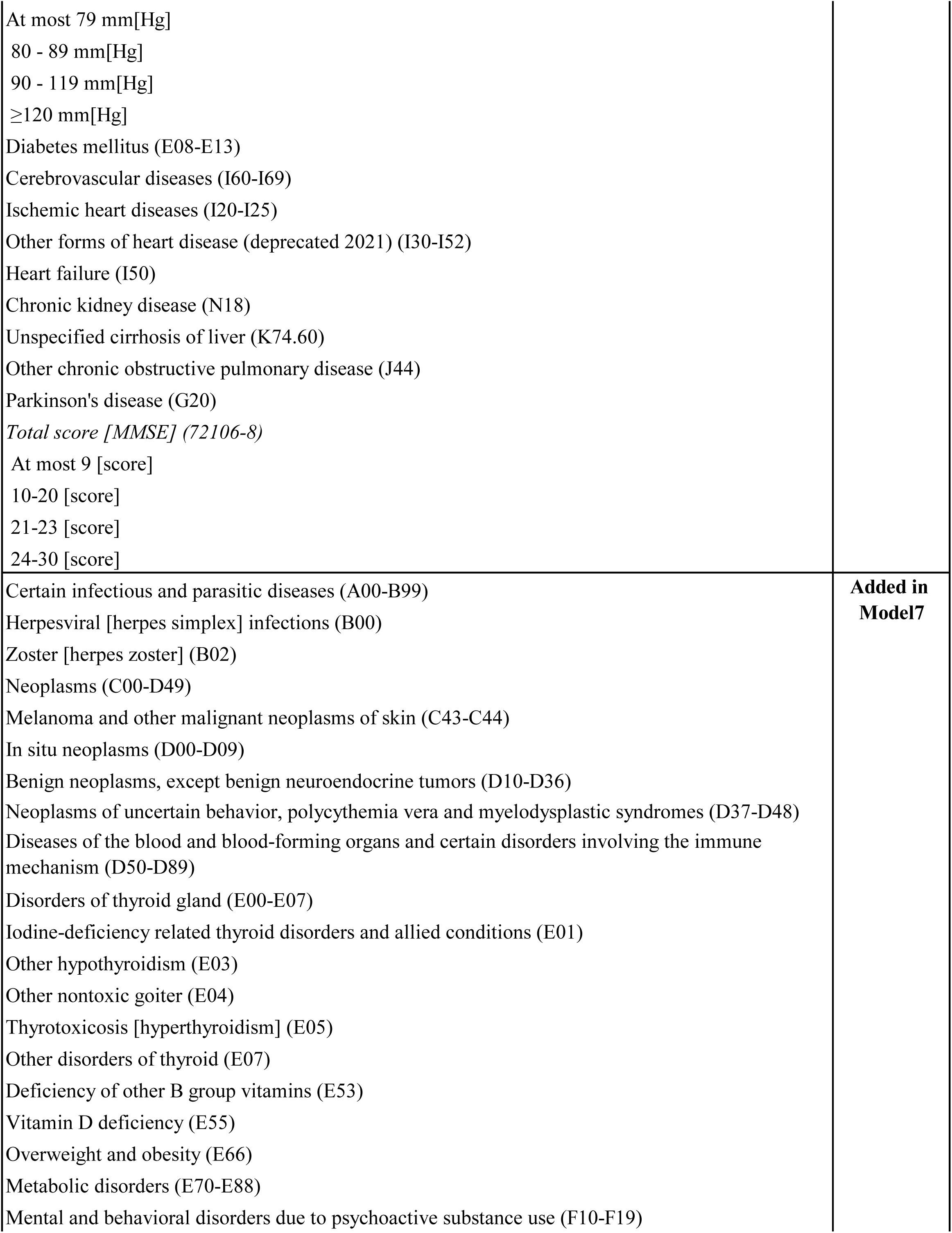
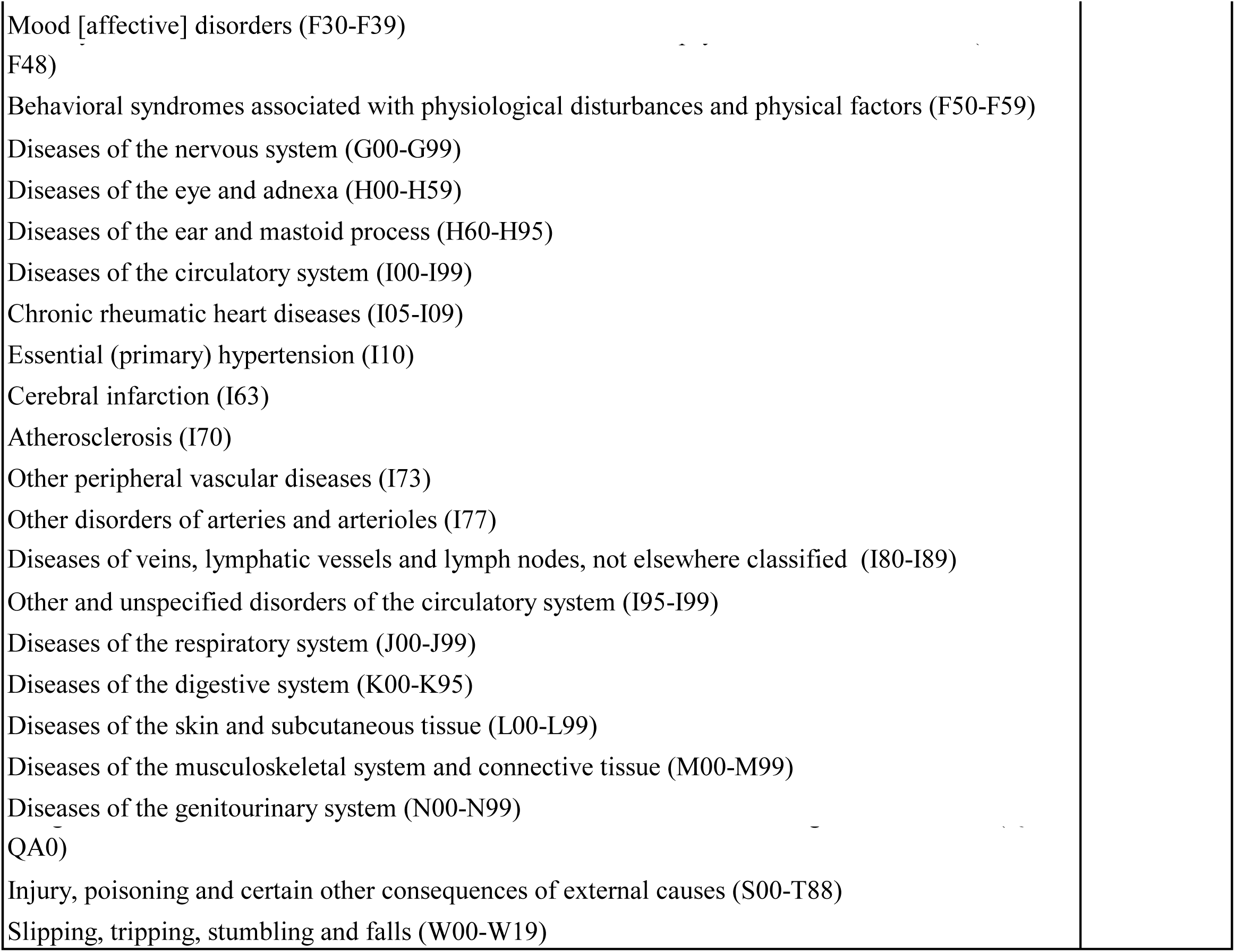
Covariates included in the sequential Cox proportional hazards models for mortality. The table summarizes the covariates added at each stage of the nested Cox proportional hazards models examining the association between Shingrix-vaccinated and non-Shingrix-vaccinated individuals and mortality in the unmatched cohorts. Model 1 was unadjusted, whereas Models 2-7 included progressively broader adjustment for sociodemographic, lifestyle, healthcare utilization, socioeconomic, clinical, and diagnostic variables. Codes in parentheses denote the corresponding TriNetX-recorded codes.

| Covariate | Model |
| --- | --- |
| No covariates | **Model 1** |
| Male (M)<br>Age at Index (AI)<br>White (2106-3)<br>Black or African American (2054-5)<br>Asian (2028-9)<br>American Indian or Alaska Native (1002-5)<br>Native Hawaiian or Other Pacific Islander (2076-8)<br>Other Race (2131-1) | **Added in Model 2** |
| *BMI (9083)*<br>At most 18.4 kg/m<sup>2</sup><br>18.5-24.9 kg/m<sup>2</sup><br>25-29.9 kg/m<sup>2</sup><br>≥ 30 kg/m<sup>2</sup><br>Tobacco use (Z72.0)<br>Personal history of nicotine dependence (Z87.891)<br>Alcohol abuse (F10.1) | **Added in Model 3** |
| Encounter for general adult medical examination without abnormal findings (Z00.00)<br>Encounter for general adult medical examination with abnormal findings (Z00.01)<br>Long term (current) drug therapy (Z79)<br>Encounter for screening for malignant neoplasms (Z12) | **Added in Model 4** |
| Problems related to employment and unemployment (Z56)<br>Problems related to housing and economic circumstances (Z59)<br>T: State Medicaid Agency Codes (T)<br>S: Private Payer Codes (S) | **Added in Model 5** |
| *Blood Pressure, Systolic (9085)*<br>At most 119 mm[Hg]<br>120 - 129 mm[Hg]<br>130 - 139 mm[Hg]<br>140 - 179 mm[Hg]<br>≥180 mm[Hg]<br>*Blood Pressure, Diastolic (9086)* | **Added in Model 6** |
| At most 79 mm[Hg]<br>80 - 89 mm[Hg]<br>90 - 119 mm[Hg]<br>≥120 mm[Hg]<br>Diabetes mellitus (E08-E13)<br>Cerebrovascular diseases (I60-I69)<br>Ischemic heart diseases (I20-I25)<br>Other forms of heart disease (deprecated 2021) (I30-I52)<br>Heart failure (I50)<br>Chronic kidney disease (N18)<br>Unspecified cirrhosis of liver (K74.60)<br>Other chronic obstructive pulmonary disease (J44)<br>Parkinson's disease (G20)<br>*Total score [MMSE] (72106-8)*<br>At most 9 [score]<br>10-20 [score]<br>21-23 [score]<br>24-30 [score] |  |
| Certain infectious and parasitic diseases (A00-B99)<br>Herpesviral [herpes simplex] infections (B00)<br>Zoster [herpes zoster] (B02)<br>Neoplasms (C00-D49)<br>Melanoma and other malignant neoplasms of skin (C43-C44)<br>In situ neoplasms (D00-D09)<br>Benign neoplasms, except benign neuroendocrine tumors (D10-D36)<br>Neoplasms of uncertain behavior, polycythemia vera and myelodysplastic syndromes (D37-D48)<br>Diseases of the blood and blood-forming organs and certain disorders involving the immune mechanism (D50-D89)<br>Disorders of thyroid gland (E00-E07)<br>Iodine-deficiency related thyroid disorders and allied conditions (E01)<br>Other hypothyroidism (E03)<br>Other nontoxic goiter (E04)<br>Thyrotoxicosis [hyperthyroidism] (E05)<br>Other disorders of thyroid (E07)<br>Deficiency of other B group vitamins (E53)<br>Vitamin D deficiency (E55)<br>Overweight and obesity (E66)<br>Metabolic disorders (E70-E88)<br>Mental and behavioral disorders due to psychoactive substance use (F10-F19) | **Added in Model7** |
| Mood [affective] disorders (F30-F39)<br>F48)<br>Behavioral syndromes associated with physiological disturbances and physical factors (F50-F59)<br>Diseases of the nervous system (G00-G99)<br>Diseases of the eye and adnexa (H00-H59)<br>Diseases of the ear and mastoid process (H60-H95)<br>Diseases of the circulatory system (I00-I99)<br>Chronic rheumatic heart diseases (I05-I09)<br>Essential (primary) hypertension (I10)<br>Cerebral infarction (I63)<br>Atherosclerosis (I70)<br>Other peripheral vascular diseases (I73)<br>Other disorders of arteries and arterioles (I77)<br>Diseases of veins, lymphatic vessels and lymph nodes, not elsewhere classified (I80-I89)<br>Other and unspecified disorders of the circulatory system (I95-I99)<br>Diseases of the respiratory system (J00-J99)<br>Diseases of the digestive system (K00-K95)<br>Diseases of the skin and subcutaneous tissue (L00-L99)<br>Diseases of the musculoskeletal system and connective tissue (M00-M99)<br>Diseases of the genitourinary system (N00-N99)<br>QA0)<br>Injury, poisoning and certain other consequences of external causes (S00-T88)<br>Slipping, tripping, stumbling and falls (W00-W19) |  |

## References

1. Li J, Li J. Trends, inequalities, and cross-location similarities in global dementia burden and attributable risk factors across 204 countries and territories: a systematic analysis for the Global Burden of Disease Study 2021. Int J Surg. 2025;111(8):5298–5310. doi:10.1097/JS9.0000000000002628

2. Duan Y, Han C, Zheng H, Yu J, Luo M. Global, regional, and national burden of Alzheimer’s disease and other dementias from 1990 to 2021: findings from the Global Burden of Disease Study 2021. Front Aging Neurosci. 2025;17:1678212. doi:10.3389/fnagi.2025.1678212

3. Aljuhani M, Ashraf A, Edison P. Evaluating clinical meaningfulness of anti-β-amyloid therapies amidst amyloid-related imaging abnormalities concern in Alzheimer’s disease. Brain Commun. 2024;6(6):fcae435. doi:10.1093/braincomms/fcae435

4. Alkhalifa AE, Al Mokhlf A, Ali H, Al-Ghraiybah NF, Syropoulou V. Anti-Amyloid Monoclonal Antibodies for Alzheimer’s Disease: Evidence, ARIA Risk, and Precision Patient Selection. J Pers Med. 2025;15(9):437. doi:10.3390/jpm15090437

5. Huang X, Gu BJ. Shingles vaccination and neuroimmune vulnerability. Trends Neurosci. 2025;48(9):655-662. doi:10.1016/j.tins.2025.07.003

6. Azmal M, Paul JK, Prima FS, Haque ANMSNB, Meem M, Ghosh A. Microglial dysfunction in Alzheimer’s disease: Mechanisms, emerging therapies, and future directions. Exp Neurol. 2025;392:115374. doi:10.1016/j.expneurol.2025.115374

7. Chaudhary V, Singh AP, Sharma H, Taumar D. The Role of Microglial Cells and Cytokine Modulation in Alzheimer’s Disease: A Neuroinflammatory Perspective. Curr Alzheimer Res. Published online January 2, 2026. doi:10.2174/0115672050412711251115023127

8. Mary A, Mancuso R, Heneka MT. Immune Activation in Alzheimer Disease. Annu Rev Immunol. 2024;42(1):585–613. doi:10.1146/annurev-immunol-101921-035222

9. Wainberg M, Luquez T, Koelle DM, et al. The viral hypothesis: how herpesviruses may contribute to Alzheimer’s disease. Mol Psychiatry. 2021;26(10):5476–5480. doi:10.1038/s41380-021-01138-6

10. Devanand DP. Viral Hypothesis and Antiviral Treatment in Alzheimer’s Disease. Curr Neurol Neurosci Rep. 2018;18(9):55. doi:10.1007/s11910-018-0863-1

11. Moir RD, Lathe R, Tanzi RE. The antimicrobial protection hypothesis of Alzheimer’s disease. Alzheimers Dement J Alzheimers Assoc. 2018;14(12):1602–1614. doi:10.1016/j.jalz.2018.06.3040

12. Itzhaki RF, Lathe R, Balin BJ, et al. Microbes and Alzheimer’s Disease. J Alzheimers Dis JAD. 2016;51(4):979-984. doi:10.3233/JAD-160152

13. Gilden D, Cohrs RJ, Mahalingam R, Nagel MA. Varicella zoster virus vasculopathies: diverse clinical manifestations, laboratory features, pathogenesis, and treatment. Lancet Neurol. 2009;8(8):731–740. doi:10.1016/S1474-4422(09)70134-6

14. Livieratos A, Schiro LE, Gogos C, Ntaios G, Akinosoglou K. Varicella Zoster Virus and Stroke: An Intricate Relationship. Viruses. 2025;17(12):1591. doi:10.3390/v17121591

15. Polisky V, Littmann M, Triastcyn A, et al. Varicella-zoster virus reactivation and the risk of dementia. Nat Med. 2025;31(12):4172–4179. doi:10.1038/s41591-025-03972-5

16. Shah S, Dahal K, Thapa S, et al. Herpes zoster vaccination and the risk of dementia: A systematic review and meta-analysis. Brain Behav. 2024;14(2):e3415. doi:10.1002/brb3.3415

17. Eyting M, Xie M, Michalik F, Heß S, Chung S, Geldsetzer P. A natural experiment on the effect of herpes zoster vaccination on dementia. Nature. 2025;641(8062):438–446. doi:10.1038/s41586-025-08800-x

18. Maggi S, Fulöp T, De Vita E, et al. Association between vaccinations and risk of dementia: a systematic review and meta-analysis. Age Ageing. 2025;54(11):afaf331. doi:10.1093/ageing/afaf331

19. Greenblatt CL, Lathe R. Vaccines and Dementia: Part I. Non-Specific Immune Boosting with BCG: History, Ligands, and Receptors. J Alzheimers Dis JAD. 2024;98(2):343–360. doi:10.3233/JAD-231315

20. Greenblatt CL, Lathe R. Vaccines and Dementia: Part II. Efficacy of BCG and Other Vaccines Against Dementia. J Alzheimers Dis JAD. 2024;98(2):361–372. doi:10.3233/JAD-231323

21. Taquet M, Todd JA, Harrison PJ. Lower risk of dementia with AS01-adjuvanted vaccination against shingles and respiratory syncytial virus infections. NPJ Vaccines. 2025;10(1):130. doi:10.1038/s41541-025-01172-3

22. Taquet M, Dercon Q, Todd JA, Harrison PJ. The recombinant shingles vaccine is associated with lower risk of dementia. Nat Med. 2024;30(10):2777–2781. doi:10.1038/s41591-024-03201-5

23. Xie M, Eyting M, Bommer C, Ahmed H, Geldsetzer P. The effect of shingles vaccination at different stages of the dementia disease course. Cell. 2025;188(25):7049–7064.e20. doi:10.1016/j.cell.2025.11.007

24. Cunningham AL, Lal H, Kovac M, et al. Efficacy of the Herpes Zoster Subunit Vaccine in Adults 70 Years of Age or Older. N Engl J Med. 2016;375(11):1019–1032. doi:10.1056/NEJMoa1603800

25. Oxman MN, Levin MJ, Johnson GR, et al. A vaccine to prevent herpes zoster and postherpetic neuralgia in older adults. N Engl J Med. 2005;352(22):2271–2284. doi:10.1056/NEJMoa051016

26. Cunningham AL, Lal H, Kovac M, et al. Efficacy of the Herpes Zoster Subunit Vaccine in Adults 70 Years of Age or Older. N Engl J Med. 2016;375(11):1019–1032. doi:10.1056/NEJMoa1603800

27. Greenberg GM, Koshy PA, Hanson MJS. Adult Vaccination. Am Fam Physician. 2022;106(5):534-542. Accessed March 11, 2026. https://www.aafp.org/pubs/afp/issues/2022/1100/adult-vaccination.html

28. Arevalo-Rodriguez I, Smailagic N, Roqué I Figuls M, et al. Mini-Mental State Examination (MMSE) for the detection of Alzheimer’s disease and other dementias in people with mild cognitive impairment (MCI). Cochrane Database Syst Rev. 2015;2015(3):CD010783. doi:10.1002/14651858.CD010783.pub2

29. Dow CT, Greenblatt CL, Chan ED, Dow JF. Evaluation of BCG Vaccination and Plasma Amyloid: A Prospective, Pilot Study with Implications for Alzheimer’s Disease. Microorganisms. 2022;10(2):424. doi:10.3390/microorganisms10020424

30. Greenblatt CL, Bercovier H, Klein BY, Gofrit ON. Intravesical Bacille Calmette-Guerin (BCG) Vaccine Affects Cognition. J Alzheimers Dis JAD. 2024;100(3):771–774. doi:10.3233/JAD-240307

31. Netea MG, Joosten LAB, Latz E, et al. Trained immunity: A program of innate immune memory in health and disease. Science. 2016;352(6284):aaf1098. doi:10.1126/science.aaf1098

32. Koeken VA, de Bree LCJ, Mourits VP, et al. BCG vaccination in humans inhibits systemic inflammation in a sex-dependent manner. J Clin Invest. 2020;130(10):5591–5602. doi:10.1172/JCI133935

33. Keefe RC, Takahashi H, Tran L, et al. BCG therapy is associated with long-term, durable induction of Treg signature genes by epigenetic modulation. Sci Rep. 2021;11(1):14933. doi:10.1038/s41598-021-94529-2

34. Wilkinson T, Ly A, Schnier C, et al. Identifying dementia cases with routinely collected health data: A systematic review. Alzheimers Dement. 2018;14(8):1038–1051. doi:10.1016/j.jalz.2018.02.016

35. Johnson AH, Brennan JC, King PJ, Turcotte JJ, MacDonald JH. Comparison of Postoperative Outcomes of Patients Undergoing Total Hip and Total Knee Arthroplasty Following a Diagnosis of Dementia: A TriNetX Database Study. Arthroplasty Today. 2024;27:101359. doi:10.1016/j.artd.2024.101359

36. Austin PC. Balance diagnostics for comparing the distribution of baseline covariates between treatment groups in propensity-score matched samples. Stat Med. 2009;28(25):3083–3107. doi:10.1002/sim.3697

